# Acute liver injury and its association with death risk of patients with COVID-19: a hospital-based prospective case-cohort study

**DOI:** 10.1101/2020.04.02.20050997

**Authors:** Lin Fu, Jun Fei, Shen Xu, Hui-Xian Xiang, Ying Xiang, Zhu-Xia Tan, Meng-Die Li, Fang-Fang Liu, Ying Li, Ming-Feng Han, Xiu-Yong Li, Hui Zhao, De-Xiang Xu

**Affiliations:** Second Affiliated Hospital, Anhui Medical University, Hefei, Anhui Province, 230032, China; School of Public Health, Anhui Medical University, Hefei, Anhui Province, 230032, China; Union Hospital, Huazhong University of Science and Technology, Wuhan, Hubei Province, 430040, China; The second People’s Hospital of Fuyang City, Fuyang, Anhui Province, 236015, China

**Keywords:** Severe acute respiratory syndrome coronavirus-2 (SARS-CoV-2), coronavirus disease 2019 (COVID-19), acute liver injury (ALI)

## Abstract

**Background:** Coronavirus disease 2019 (COVID-19) is a newly respiratory infectious disease caused by severe acute respiratory syndrome coronavirus-2 (SARS-CoV-2) with multiple organ injuries. The aim of this study was to analyze SARS-CoV-2-induced acute liver injury (ALI), its association with death risk and prognosis after discharge.

**Methods:** Three-hundred and fifty-five COVID-19 patients were recruited. Clinical data were collected from electronic medical records. ALI was evaluated and its prognosis was tracked. The association between ALI and death risk was analyzed.

**Results:** Of 355 COVID-19 patients, 211 were common, 88 severe, and 51 critical ill cases, respectively. On admission, 223 (62.8%) patients were with hypoproteinemia, 151(42.5%) with cholestasis, and 101 (28.5%) with hepatocellular injury. As expected, ALI was more common in critical ill patients. By multivariate logistic regression, male, older age and lymphocyte reduction were three important independent risk factors predicting ALI among COVID-19 patients. Death risk analysis shows that fatality rate was higher among patients with hypoproteinemia than those without hypoproteinemia (*RR*=9.471, *P*<0.001). Moreover, fatality rate was higher among patients with cholestasis than those without cholestasis (*RR*=2.182, *P*<0.05). Follow-up observation found that more than one hepatic functional indexes of two-third patients remained abnormal 14 days after discharge.

**Conclusions:** ALI at early stage elevates death risk of COVID-19 patients. SARS-CoV-2-induced ALI has not recovered completely 14 days after discharge.

## 1 Introduction

Since December 2019, a novel coronavirus-induced respiratory infectious disease was found in Wuhan city of central China [1]. This novel coronavirus was named as severe acute respiratory syndrome coronavirus-2 (SARS-CoV-2) by the International Committee on Taxonomy of Viruses. SARS-CoV-2-induced disease was named as coronavirus disease 2019 (COVID-19) by the World Health Organization (WHO). Since 2020, COVID-19 has been rapidly pandemic all over the world [2]. Until 31 March, 2020, more than 700,000 cases were infected with SARS-CoV-2. Surprisingly, more than 33000 patients died from COVID-19 all over the world [1].

SARS-CoV-2 is mainly transmitted by droplets or direct contact and infected through respiratory tract [3]. In addition, SARS-CoV-2 can be transmitted by feces [4]. Recently, angiotensin-converting enzyme 2 (ACE2), expressed mainly in pulmonary epithelial cells, and probably cardiomyocytes and renal tubular epithelial cells, has been identified as functional receptor for SARS-CoV-2 [5-7]. The main symptoms and signs of COVID-19 patients include fever, companied by dry cough, dyspnea, diarrhea, fatigue and lymphopenia [8-13]. Although only a few cases died in mild COVID-19 patients, death risk was abruptly increased among critical ill patients with the fatality rate even more than 50% [14]. A large number of clinical data revealed that infection with SARS-CoV-2 not only caused severe acute respiratory syndrome but also multiple organ injuries, including lymphocyte reduction, myocardial dysfunction and even acute renal failure [15-17]. Nevertheless, the clinical characteristics of acute liver injury (ALI) caused by SARS-CoV-2 are rarely described. In addition, the clinical significance of SARS-CoV-2-induced ALI and its prognosis need to be clarified.

The objective of the present study was to analyze SARS-CoV-2-induced ALI, its association with death risk and prognosis after discharge. Our results showed that male elderly COVID-19 patients with lymphopenia were more susceptible to ALI. We found that ALI at the early stage elevated death of COVID-19 patients. We provide the first evidence that SARS-CoV-2-induced ALI has not recovered completely 14 days after discharge.

## 2 Methods

### 2.1 Study design and participants

In the present study, 200 patients, who were confirmed to be infected with SARS-CoV-2 using real-time RT-PCR, were recruited from Union Hospital of Huazhong University of Science and Technology in Wuhan city. Another 155 patients, who were confirmed to be infected with SARS-CoV-2 using real-time RT-PCR, were recruited from the Second People’s Hospital of Fuyang City in Anhui province. The Second Affiliated Hospital of Anhui Medical University sent a medical team to the Union Hospital of Huazhong University of Science and Technology to recuse COVID-19 patients. The Second Affiliated Hospital of Anhui Medical University sent a specialist group to tthe Second People’s Hospital of Fuyang City to guide COVID-19 treatment. All COVID-19 patients were clinically diagnosed on basis of typical clinical manifestations accompanied with characteristic chest radiology changes. No patients with COVID-19 died in the Second People’s Hospital of Fuyang City. All patients were cured and discharged. At last, 150 cured patients with COVID-19 were performed follow-up examinations 14 days after discharge in the Second People’s Hospital of Fuyang City, biochemical indexes and blood routine among patients were detected again. All COVID-19 patients were eligible in this study. Each COVID-19 patient gave advanced oral consent. The present study was approved by the Ethics Committee of Anhui Medical University (2020-5).

### 2.2 Data collection

The medical record of each COVID-19 patient was evaluated. Following data were collected from the electronic medical records: demographic information, preexisting comorbidities, including chronic obstructive pulmonary disease, hepatic disease, cardiovascular disease, hypertension, diabetes and other disease. Patient’s signs and symptoms, and laboratory test results were also collected. The dates of onset, admission and death were recorded. The onset time was defined as the date when patients’ any symptom and sign were found.

### 2.3 Laboratory testing

Patient’s pharyngeal swab specimens were collected for extraction of SARS-CoV-2 RNA. Real-time RT-PCR was used to detect viral nucleic acid using a COVID-19 nucleic acid detection kit following experimental instructions (Shanghai bio-germ Medical Technology Co Ltd). Viral RNA extraction and nucleic acid detection were executed by either the Center for Disease Control and Prevention of Wuhan City or the Center for Disease Control and Prevention of Fuyang City. Hepatic function indexes included: Alanine aminotransferase (ALT), aspartate aminotransferase (AST), AST/ALT ratio, total bilirubin (TBIL), direct bilirubin (DBIL), indirect bilirubin (IBIL), total protein, albumin, globulin, albumin/globulin ratio, alkaline phosphatase (ALP), glutamyl transferase (γ-GGT), total bile acid (TBA), and oxygenation index were examined on admission. All laboratory tests were analyzed by the clinical laboratory of either the Union Hospital of Huazhong University of Science and Technology or the Second People’s Hospital of Fuyang City.

### 2.4 Statistical analysis

All statistical analyses were performed using SPSS 21.0 software. Categorical variables were expressed with frequencies and percentages. Continuous variables were shown using median and mean values. Means for continuous variables were compared with independent-samples *t* tests when the data were normally distributed; if not, the Mann-Whitney test was used. Proportions for categorical variables were compared with the *chi-square* and *Fisher’s* exact test. Univariable logistic regression between basic disease or different parameter and demise was performed. Moreover, the main risks related with demise were examined using multivariable logistic regression models adjusted for potential confounders. Statistical significance was determined at *P*<0.05.

## 3 Results

### 3.1 Demographic and clinical characteristics of COVID-19 patients

The clinical characteristics of 350 COVID-19 patients were analyzed. As shown in Table 1, common case, defined as oxygenation index higher than 300, was 60.3%. For severe case, whose oxygenation index was from 200 to 300, was 25.1%. For critically ill case, whose oxygenation index was lower than 200, was 14.6% (Table 1). The demographic characteristics of 350 COVID-19 patients were then analyzed. Of 350 COVID-19 patients, males accounted for 53.5% and females accounted for 46.5% (Table 2). There were 103 patients younger than 40, 137 cases aged between 40 and 60, and 115 patients older than 60 (Table 2). As shown in Table 2, 145 patients were with diabetes, 125 with hypertension and 16 with hepatic diseases. Finally, blood lymphocytes were analyzed among COVID-19 patients. As shown in Table 2, 61.1% (217/355) patients were with lymphopenia.

**Table 1.** The association between the severity and hepatic function markers among COVID-19 patients.

|  | Common patients<br>(N=211) | Severe patients<br>(N=88) | Critical ill patients<br>(N=51) |
| --- | --- | --- | --- |
| Hepatocellular injury markers |  |  |  |
| Total bilirubin (μmol/L) | 10.4 (7.5, 14.7) | 10.9 (8.0, 16.2) | 12.6 (10.5, 17.0) **<sup>¶</sup> |
| Direct bilirubin (μmol/L) | 3.4 (2.6, 4.9) | 3.6 (2.4, 5.3) | 4.3 (3.0, 6.1) * |
| Indirect bilirubin (μmol/L) | 7.0 (4.7, 10.0) | 7.6 (5.5, 11.0) | 8.3 (6.2, 10.8) ** |
| ALT (U/L) | 22.0 (14.0, 35.0) | 23.0 (15.0, 42.0) | 33.0 (19.0, 61.0) ** |
| AST (U/L) | 26.0 (20.0, 35.0) | 29.0 (23.0, 54.0)** | 49.0 (35.0, 80.0) **<sup>¶¶</sup> |
| AST/ALT ratio | 1.2 (0.9, 1.7) | 1.3 (1.0, 1.8) | 1.6 (1.1, 2.2) **<sup>¶</sup> |
| Hypoproteinemia markers |  |  |  |
| Total protein (g/L) | 69.9 (64.8, 74.8) | 65.0 (59.1, 70.1)** | 61.0 (58.8, 65.3) **<sup>¶¶</sup> |
| Albumin (g/L) | 40.5 (36.8, 44.0) | 37.3 (33.7, 40.0) ** | 34.5 (31.7, 37.4) **<sup>¶¶</sup> |
| Globulin (g/L) | 29.4 (25.9, 32.9) | 27.9 (23.8, 31.4)* | 27.3 (23.6, 30.1) |
| Aibumin/globulin ratio | 1.4 (1.2, 1.5) | 1.3 (1.2, 1.7) | 1.2 (1.0, 1.5) **<sup>¶</sup> |
| Cholestasis markers |  |  |  |
| Alkaline phosphatase (U/L) | 62.0 (51.0, 77.0) | 62.0 (51.0, 71.0) | 71.5 (57.8, 102.0) **<sup>¶¶</sup> |
| Glutamyl transferase (U/L) | 24.0 (15.0, 41.0) | 26.0 (18.0, 51.8) | 42.5 (26.5, 93.2) **<sup>¶¶</sup> |
| TBA (μmol/L) | 4.8 (3.0, 7.0) | 4.0 (2.5, 7.7) | 4.0 (2.2, 5.8) |

Five patients' information of the severity was missed.
Compared with common patients, \* $P<0.05$ , \*\* $P<0.01$ ;
Compared with severe patients, <sup>¶</sup> $P<0.05$ , <sup>¶¶</sup> $P<0.01$ .

**Table 2.** The effects of demographic characteristics and complications on hepatic function among COVID-19 patients.

|  | Cases, N | DBIL (μmol/L) | IBIL (μmol/L) | ALT (U/L) | Albumin (g/L) | Globulin (g/L) | ALP (U/L) | γ-GGT (U/L) | TBA (μmol/L) |
| --- | --- | --- | --- | --- | --- | --- | --- | --- | --- |
| Gender |  |  |  |  |  |  |  |  |  |
| Male | 190 | 4.2 (3.1, 5.9) | 8.1 (5.9, 11.6) | 29.0 (18.0, 49.8) | 39.0 (35.1, 43.1) | 28.3 (24.7, 32.0) | 64.0 (53.0, 78.0) | 33.0 (21.0, 70.0) | 4.8 (3.0, 7.4) |
| Female | 165 | 2.9 (2.2, 4.1) ** | 6.5 (4.4, 8.7) ** | 19.0 (13.0, 28.0) ** | 38.6 (34.3, 41.3) ** | 28.7 (25.7, 32.5) | 61.0 (49.0, 77.0) ** | 20.0 (13.0, 35.3) ** | 4.0 (2.8, 6.9) |
| Age (year) |  |  |  |  |  |  |  |  |  |
| < 40 | 103 | 3.4 (2.6, 4.6) | 6.2 (4.2, 9.3) | 12.0 (13.0, 38.0) | 42.0 (38.8, 45.8) | 29.6 (26.3, 32.9) | 58.0 (47.3, 69.8) | 20.5 (11.0, 36.0) | 4.8 (3.3, 6.9) |
| 41-59 | 137 | 3.6 (2.4, 5.4) | 7.6 (5.5, 10.6) | 24.0 (16.0, 42.0) ** | 38.8 (35.5, 41.8) ** | 28.8 (25.2, 32.3) | 62.0 (52.0, 75.0) * | 30.0 (19.0, 69.0) ** | 4.8 (3.0, 8.0) |
| > 60 | 115 | 4.0 (2.7, 5.4) ** | 8.3 (6.5, 11.3) ** | 25.0 (17.5, 35.0) ** | 35.3 (32.3, 39.5) **¶ | 27.1 (23.2, 31.4) ** | 67.0 (55.0, 80.0) ** | 28.0 (18.0, 48.7) ** | 3.9 (2.0, 5.9) *¶ |
| ¶ |  |  |  |  |  |  |  |  |  |
| Hypertension |  |  |  |  |  |  |  |  |  |
| Yes | 125 | 3.6 (2.6, 5.0) | 7.8 (5.8, 10.3) | 24.0 (14.0, 42.0) | 37.6 (33.4, 40.4) | 27.2 (24.1, 30.3) | 63.5 (54.0, 82.8) | 28.0 (18.0, 63.0) | 4.2 (2.6, 7.0) |
| No | 230 | 3.6 (2.6, 5.0) | 7.3 (4.8, 10.6) | 23.0 (16.0, 38.0) | 39.8 (35.7, 43.3) ** | 29.2 (25.5, 32.8) ** | 62.0 (50.5, 75.0) * | 26.0 (16.3, 46.0) | 4.4 (2.9, 7.0) |
| Diabetes |  |  |  |  |  |  |  |  |  |
| Yes | 145 | 11.4 (8.2, 15.6) | 3.5 (2.2, 5.1) | 22.5 (15.0, 42.0) | 37.3 (33.3, 40.6) | 26.5 (22.9, 29.4) | 65.0 (54.0, 86.0) | 28.5 (19.0, 63.8) | 4.4 (2.5, 7.0) |
| No | 210 | 10.7 (7.8, 15.1) | 3.7 (2.7, 5.0) | 23.0 (15.0, 38.0) | 39.9 (36.2, 43.5) ** | 30.0 (26.9, 33.3) ** | 61.5 (50.0, 72.3) ** | 26.0 (16.0, 42.3) * | 4.4 (3.0, 7.0) |
| Hepatic diseases |  |  |  |  |  |  |  |  |  |
| Yes | 16 | 3.4 (2.4, 11.9) | 6.2 (4.2, 15.4) | 24.5 (17.8, 49.5) | 36.7 (27.2, 40.3) | 28.1 (24.2, 31.1) | 63.0 (53.0, 73.3) | 24.5 (16.0, 35.0) | 4.9 (2.5, 8.5) |
| No | 339 | 3.6 (2.6, 5.0) | 7.5 (5.3, 10.4) | 23.0 (15.0, 39.3) | 38.8 (34.8, 42.4) | 28.7 (25.0, 32.3) | 62.0 (51.0, 78.0) | 27.0 (17.0, 50.0) | 4.3 (2.8, 7.0) |
| Lymphopenia |  |  |  |  |  |  |  |  |  |
| Yes | 217 | 3.7 (2.4, 5.3) | 7.2 (5.0, 10.6) | 25.0 (16.0, 45.0) | 37.6 (34.1, 40.7) | 27.9 (24.1, 31.8) | 63.0 (51.0, 79.0) | 30.0 (18.0, 68.0) | 4.7 (3.0, 7.5) |
| No | 138 | 3.4 (2.7, 4.3) | 7.8 (5.2, 10.3) | 21.0 (13.0, 33.5) ** | 41.2 (37.5, 44.8) ** | 29.3 (26.1, 32.7) ** | 62.0 (52.5, 72.5) | 25.0 (15.5, 38.5) * | 4.2 (2.7, 6.6) |

Cases in Gender, compared with “Male” group, \* $P<0.05$ , \*\* $P<0.01$ .
Cases in Age, compared with “< 40” group, \* $P<0.05$ , \*\* $P<0.01$ ; Compared with “41-59” group, <sup>†</sup> $P<0.05$ , <sup>††</sup> $P<0.01$ .
Cases in Hypertension, compared with “Yes” group, \* $P<0.05$ , \*\* $P<0.01$ .
Cases in Diabetes, compared with “Yes” group, \* $P<0.05$ , \*\* $P<0.01$ .
Cases in Lymphocyte reduction, compared with “Yes” group, \* $P<0.05$ , \*\* $P<0.01$ .

### 3.2 Association between the severity and ALI among COVID-19 patients

The association between the severity and ALI was analyzed among COVID-19 patients. As shown in Table 1, all hepatocellular injury markers, including total bilirubin, direct bilirubin, indirect bilirubin, ALT and AST, were higher in critically ill patients than those of common cases. By contrast, total protein, albumin and albumin/globulin ratio, three markers of hypoproteinemia were lower in critically ill patients than those of common cases. Despite of no difference on serum TBA, alkaline phosphatase and glutamyl transferase, two markers of cholestasis, were higher in critically ill patients than those of common cases. The association between oxygenation index and hepatic functional indexes was analyzed. As shown in Figure 1, there was a weak negative correlation between oxygenation index and ALT, AST, AST/ALT, and glutamyl transferase. By contrast, there was a weak positive correlation between oxygenation index and albumin among COVID-19 patients (Figure 1).

**Figure 1.** The association between oxygenation index and hepatic functional indices among COVID-19 patients. (A-E) Correlation between oxygenation index and hepatocellular injury indices was analyzed among COVID-19 patients. (A) Serum ALT; (B) AST; (C) AST/ALT ratio; (D) TBIL; (E) DBIL. (F-I) Correlation between oxygenation index and hypoproteinemia indices was analyzed among COVID-19 patients. (F) Serum total protein; (G) Albumin; (H) Globulin; (I) Albumin/ Globulin ratio. (J-L) Correlation between oxygenation index and cholestasis indices was analyzed among all COVID-19 patients. (J) Serum alkaline phosphatase; (K) Glutamyl transferase; (L) TBA.

### 3.3 Male elderly COVID-19 patients with lymphopenia are more susceptible to ALI

The effects of demographic characteristics on hepatic functional indexes were analyzed. As shown in Table 2, the levels of direct bilirubin, indirect bilirubin, ALT, alkaline phosphatase and glutamyl transferase were higher in males than in females. By contrast, the level of albumin was lower in males than in females. Further analysis showed that the levels of total bilirubin, ALT, alkaline phosphatase and glutamyl transferase were higher in patients older than 60 than those of younger patients. By contrast, the level of albumin was lower patients older than 60 than those of younger patients (Table 2). The effects of comorbidity on hepatic functional indexes were then analyzed. As shown in Table 2, alkaline phosphatase was slightly increased in COVID-19 patients with hypertension as compared with those without hypertension. Further analysis showed that the levels of alkaline phosphatase and glutamyl transferase were slightly increased in COVID-19 patients with diabetes as compared with those without diabetes. By contrast, the level of albumin was slightly decreased in COVID-19 patients with diabetes as compared with those without diabetes. Of interest, there was no significant association between hepatic functional indexes and comorbidity with liver disease (Table 2). Finally, the association between blood lymphocytes and ALI was analyzed among COVID-19 patients. As shown in Table 2, the levels of ALT and glutamyl transferase were higher in COVID-19 patients with lymphopenia than those without lymphopenia. By contrast, the levels of albumin and globulin were lower in COVID-19 patients with lymphopenia than those without lymphopenia (Table 2). Multivariable logistic regression was used to analyze risk factors of ALI among 355 COVID-19 patients. As shown in Supplemental Table 1, the *ORs* of males were 2.884 (95% *Cl*: 1.812, 4.591; *P*<0.001) for cholestasis and 4.047 (95% *Cl*: 2.367, 6.921; *P*<0.001) for hepatocellular injury, respectively. Further analysis showed that the *ORs* of older age were 1.593 (95% *Cl*: 1.593, 1.979; *P*<0.05) for cholestasis and 3.739 (95% *Cl*: 2.078, 6.727; *P*=0.001) for hypoproteinemia, respectively. The association between blood lymphocyte count and ALI was analyzed among COVID-19 patients. The *ORs* of lymphopenia were 2.003 (95% *Cl*: 1.241, 3.233; *P*=0.004) for cholestasis, 3.218 (95% *Cl*: 1.969, 5.260; *P*=0.001) for hypoproteinemia, and 2.292 (95% *Cl*: 1.329, 3.953; *P*=0.003) for hepatocellular injury, respectively (Supplemental Table 1). The association between comorbidities and ALI was analyzed among COVID-19 patients. The *ORs* of diabetes were 1.727 (95% *Cl*: 1.014, 2.941; *P*=0.044) for cholestasis, 1.829 (95% *Cl*: 1.010, 3.312; *P*=0.046) for hypoproteinemia, respectively. No significant relationship was observed between ALI and comorbidity with hypertension and hepatic disease among COVID-19 patients (Supplemental Table 1).

### 3.4 ALI at early stage elevates death risk of COVID-19 patients

The effects of ALI at the early stage on death risk are presented in Table 3. Among 223 COVID-19 patients with hypoproteinemia, 14.3% were died. The fatality rate was higher among COVID-19 patients with hypoproteinemia than those without hypoproteinemia (14.3 % vs 1.5%; *RR*=9.471, 95% *Cl*: 2.307, 38.880; *P*<0.001). Among 151 COVID-19 patients with cholestasis, 13.9% were died. The fatality rate was higher among COVID-19 patients with cholestasis than those without cholestasis (13.9 % vs 6.4%; *RR*=2.182, 95% *Cl*: 1.129, 4.218; *P*<0.05). As shown in Table 3, there was no significant association between hepatocellular injury and death risk among COVID-19 patients.

**Table 3.**
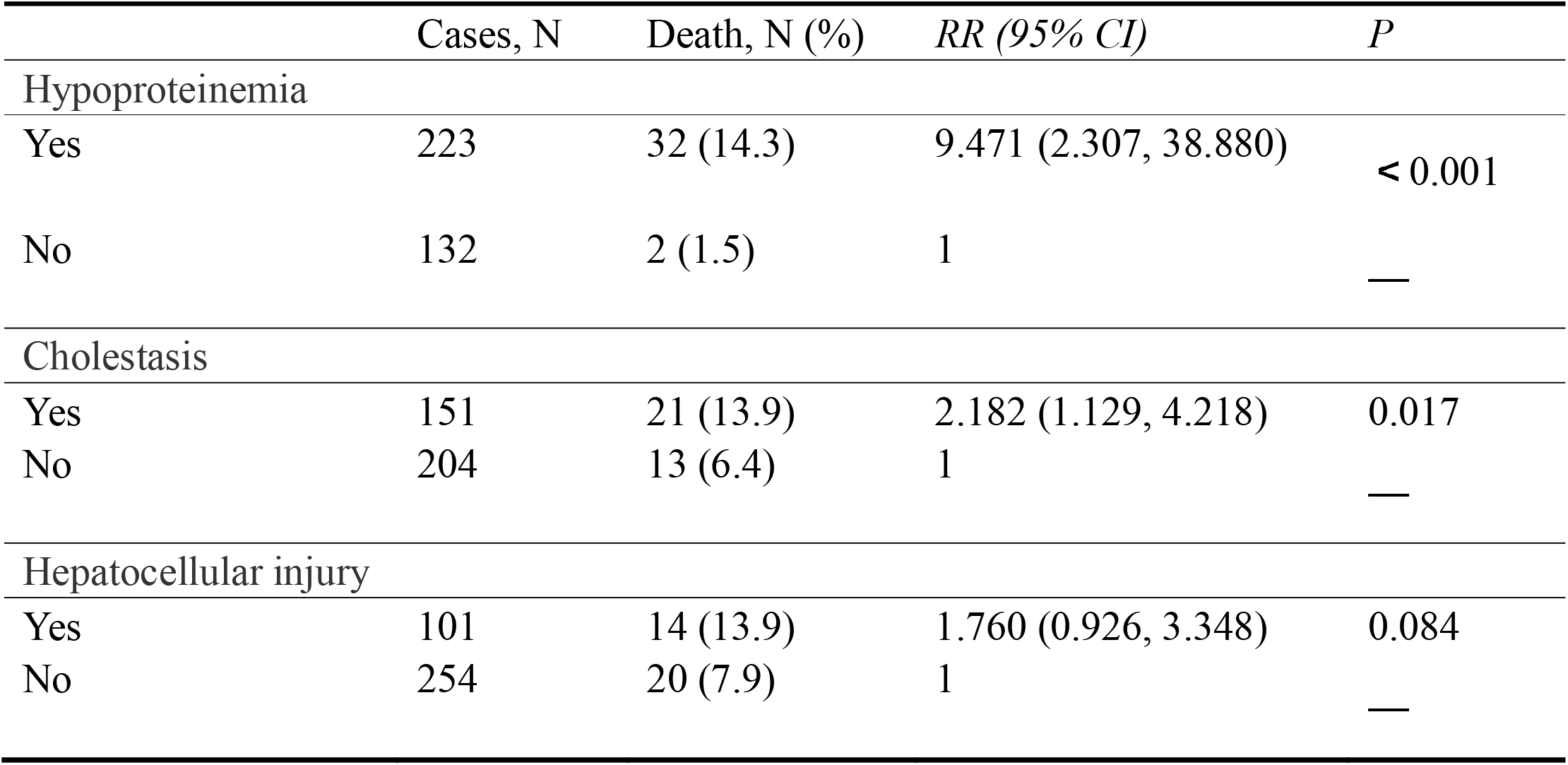
The association between ALI and death risk among COVID-19 patients.

|  | Cases, N | Death, N (%) | RR (95% CI) | P |
| --- | --- | --- | --- | --- |
| Hypoproteinemia |  |  |  |  |
| Yes | 223 | 32 (14.3) | 9.471 (2.307, 38.880) | < 0.001 |
| No | 132 | 2 (1.5) | 1 | — |
| Cholestasis |  |  |  |  |
| Yes | 151 | 21 (13.9) | 2.182 (1.129, 4.218) | 0.017 |
| No | 204 | 13 (6.4) | 1 | — |
| Hepatocellular injury |  |  |  |  |
| Yes | 101 | 14 (13.9) | 1.760 (0.926, 3.348) | 0.084 |
| No | 254 | 20 (7.9) | 1 | — |

### 3.5 Hepatic functional indexes remain abnormal 14 days after discharge

The prognosis of SARS-CoV-2-induced ALI was tracked 14 days after discharge. As shown in Table 4, no significant change was observed between hepatic functional indexes on 14 days after discharge and those on admission. Although the percentage of patients with hypoproteinemia was lower on 14 days after discharge than on admission (18.7% vs 51.9%, *P*<0.001) (Supplemental Table 2), 16.7% albumin, 1.3% pre albumin and 6.7% albumin/globulin ratio remained below normal range (Table 4). As shown in Table 4, the percentage of patients with serum ALT elevation was higher on 14 days after discharge than on admission (31.3% vs 19.5%, *P*<0.05). In addition, patients with serum IBIL elevation was higher on 14 days after discharge than on admission (4.5% vs 10.0%, *P*<0.05). No significant difference on the percentages of patients with abnormal value for direct bilirubin, TBA, alkaline phosphatase, glutamyl transferase and AST was observed when admission and 14 days after discharge (Table 4). Further analysis showed that no significant difference on the percentages of patients with cholestasis and hepatocellular injury was observed when admission and discharge, while the percentages of patients with hypoproteinemia was significantly reduced on 14 days after discharge as compared with on admission (Supplemental Table 2).

**Table 4.**
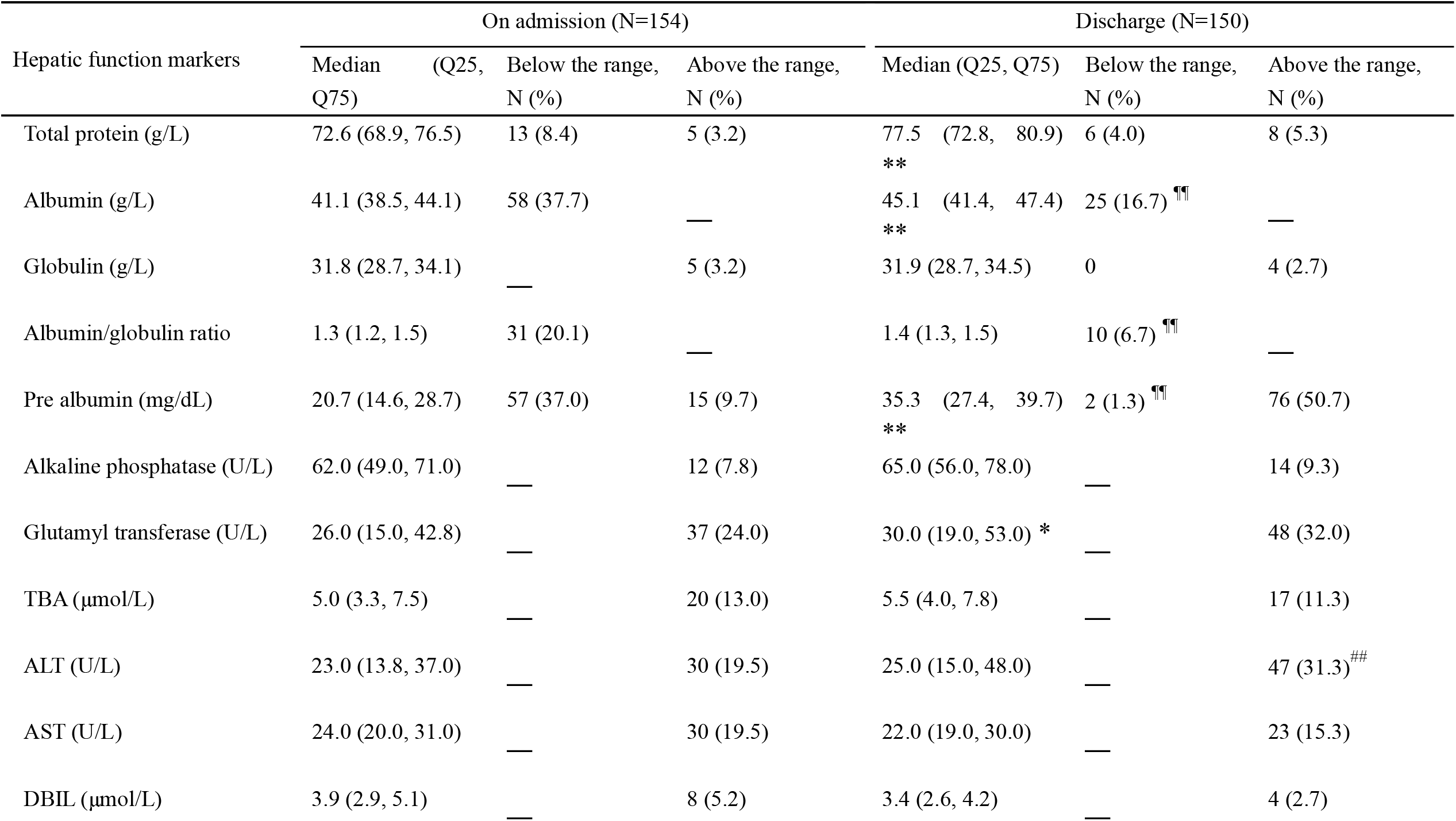
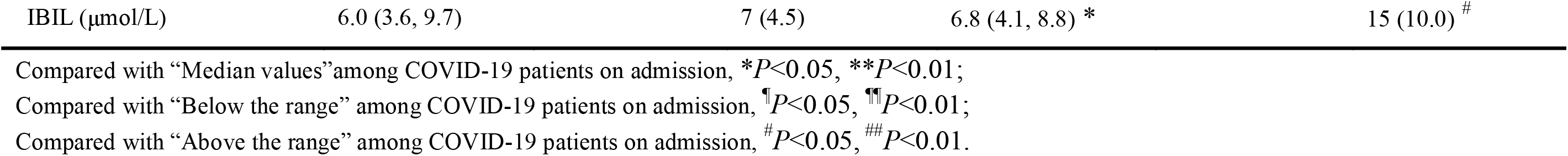
Hepatic function markers on admission and after discharge among COVID-19 patients.

## Discussion

The present study aimed to analyze SARS-CoV-2-induced ALI, its association with death risk and the prognosis after discharge. The major findings of this study include: (1) ALI is more common in the critically ill COVID-19 patients; (2) Male elderly COVID-19 patients with diabetes and lymphopenia are more susceptible to ALI; (3) ALI at the early stage elevates death risk of COVID-19 patients; (4) Hepatic functional indexes of two-third COVID-19 patients remain abnormal 14 days after discharge.

Accumulating data demonstrated that SARS-CoV-2 infection caused multiple organ injuries, including myocardial dysfunction, lymphopenia and even acute renal failure [15-17]. In the present study, we described ALI among COVID-19 patients. ALI was determined by measuring biochemical indexes, such as ALT, AST, direct bilirubin and indirect bilirubin, several hepatocellular injury markers, total protein, albumin and globulin, three protein metabolic indexes, alkaline phosphatase, TBA and glutamyl transferase, three markers of cholestasis. Our results showed that 39.6% COVID-19 patients were with cholestasis, 51.9% with hypoproteinemia, and 39.0% with hepatocellular injury on admission. We found that direct bilirubin, indirect bilirubin, ALT and AST, four indexes of hepatocellular injury, and alkaline phosphatase and glutamyl transferase, two markers of cholestasis, were higher in critically ill patients than those of common cases. By contrast, total protein and albumin were lower in the critically ill patients than those of common cases. These results provide evidence that ALI on admission is associated with the severity of COVID-19 patients.

Several studies found that elderly COVID-19 patients had more severe symptoms and signs than younger cases [18, 19]. The present study analyzed the influence of gender and age on SARS-CoV-2-induced ALI. We showed that the levels of serum total bilirubin, ALT, alkaline phosphatase and glutamyl transferase were higher in males than in females. By contrast, serum albumin level was lower in males than in females. Further analysis showed that total bilirubin, ALT, alkaline phosphatase and glutamyl transferase were higher in older patients than younger ones. By contrast, albumin and globulin were lower in older patients than younger ones. According to several clinical reports, COVID-19 patients with comorbidities had worse prognosis [14, 20]. Indeed, this study found that 40.8% COVID-19 patients were with diabetes, 35.2% with hypertension, and 4.5% with liver disease. To explore the influence of comorbidities on ALI, the present study analyzed hepatic functional indexes among different groups. Our results showed that alkaline phosphatase was slightly higher in COVID-19 patients with hypertension than those without hypertension. In addition, alkaline phosphatase and glutamyl transferase were slightly higher in patients with diabetes than those without diabetes. By contrast, albumin was slightly lower in patients with either diabetes or hypertension than those neither diabetes nor hypertension. Unexpectedly, comorbidity with liver disease did not influence hepatic functional indexes of COVID-19 patients. Lymphocytopenia is one of the important early manifestations during the pathogenesis of COVID-19 [21, 22]. This study analyzed the influence of lymphocytopenia on SARS-CoV-2-induced ALI. We found that almost all hepatic functional indexes were worse among patients with lymphocytopenia than without lymphocytopenia. To exclude potential confounding factors, multivariable logistic regression was used to further analyze the impact of gender, age and comorbidities on SARS-CoV-2-induced ALI. We found that male, older age, comorbidity with diabetes and lymphocytopenia were independent risk factors of cholestasis. In addition, older age, lymphocytopenia and comorbidity with diabetes were independent risk factors of hypoproteinemia. Moreover, male and lymphocytopenia were independent risk factors of hepatocellular injury. Taken together, the present study provide evidence that male, older age, comorbidity with diabetes and lymphocytopenia are major risk factor of ALI among COVID-19 patients.

The influence of ALI on the prognosis of COVID-19 is unclear. The present study analyzed the impact of ALI on death risk of COVID-19 patients. Our results showed that the fatality rate was higher in COVID-19 patients with hypoproteinemia than those without hypoproteinemia. Moreover, the fatality rate was higher in COVID-19 patients with cholestasis than without cholestasis. Our results suggest that hypoproteinemia and cholestasis at the early stage elevate death risk of COVID-19 patients. It is especially interesting whether SARS-CoV-2-induced ALI recovers in a short time after discharge. In the present study, 150 COVID-19 patients were followed up and measured hepatic functions 14 days after discharge. Unexpectedly, no significant difference on the values of serum direct bilirubin, indirect bilirubin, TBA, alkaline phosphatase, glutamyl transferase and AST was observed when admission and 14 days after discharge. Although serum albumin level was rebounded on 14 days after discharge, 17% COVID-19 patients remained below normal range. Our results indicate that hepatic functional indexes of two-third COVID-19 patients remain abnormal 14 days after discharge. Therefore, further follow-up is required to further evaluate whether SARS-CoV-2 infection causes permanent liver injury.

The mechanism through which SARS-CoV-2 evokes ALI remains unclear. Accumulating data indicate that SARS-CoV-2 infection causes multiple organ injuries, such as lymphopenia, myocardial dysfunction and acute renal failure [9, 15-17, 25]. In the present study, we showed that oxygenation index, an index of respiratory function, was positively correlated with serum albumin. By contrast, there was a weak negative correlation between oxygenation index and ALT, AST, AST/ALT, and glutamyl transferase among COVID-19 patients. These results suggest that respiratory failure may contribute, at least partially, to SARS-CoV-2-induced ALI. Several studies demonstrated that ACE2, as a receptor for SARS-CoV-2, was expressed in cholangiocytes and hepatocytes [26, 27]. Therefore, this study does not exclude that SARS-CoV-2 evokes ALI partially through infecting liver. It is required to further explore whether human liver is another target of SARS-CoV-2 injection.

In summary, this study aimed to investigate SARS-CoV-2-induced ALI among 355 COVID-19 patients from two hospitals. Our results revealed that SARS-CoV-2-induced ALI was more common in critically ill patients. In addition, male elderly COVID-19 patients with diabetes mellitus and lymphopenia were more susceptible to ALI. We provide evidence that ALI at the early stage elevates death risk of COVID-19 patients. Importantly, SARS-CoV-2-induced ALI has not recovered completely 14 days after discharge. Therefore, it is necessary to further evaluate whether SARS-CoV-2 infection causes permanent liver injury.

## Data Availability

All data referred to in the manuscript are available.

## AUTHOR CONTRIBUTIONS

D.X.X., H.Z. designed research; L.F., J.F., S.X., H.X.X., Y.X., Z.X.T, M.D.L, F.F.L., Y.L., M.F.H., and X.Y.L. conducted research; L.F. and J.F. analyzed data; D.X.X. and S.X. wrote the paper; D.X.X. and L.F. had primary responsibility for final content. All authors read and approved the final manuscript.

## ACKNOWLEDGMENTS

We thank all members of respiratory and critical care medicine in the second People’s Hospital of Fuyang City and Union Hospital of Huazhong University of Science and Technology for recruiting participators.

